# Epidemiological profile study of COVID-19 in West African countries: Nigeria, Senegal, Mauritania, Cape Verde and Mali

**DOI:** 10.1101/2021.05.31.21258118

**Authors:** Mouhamadou Faly Ba, Boly Diop, Oumar Bassoum, Ndèye Marème Sougou, Khadim Niang, Emmanuel Bonnet, Valéry Ridde, Adama Faye

**Author notes:** Corresponding author(s) Email address (M.F. Ba).

## Abstract

**Introduction:** The COVID-19 pandemic was first reported in West Africa on 27 February 2020 in Nigeria. It subsequently spread to other countries in the region. The objective of this study is to analyze the epidemiological profile of COVID-19 in West Africa from the first reported case to 31 January 2021.

**Method:** We publicly used available data from reliable sources and from the “COVID-19R” package. We used epidemic curves to describe the trends in the daily evolution of confirmed cases and deaths of COVID-19 in West Africa and specifically in the five countries. The reproduction rate and evolution rates were calculated from these trends.

**Results:** As of 31 January 2021, West Africa had 342,938 confirmed cases of COVID-19 with 4,496 deaths. Nigeria had 131,242 cases with 1,586 deaths. Senegal had 26,523 cases with 628 deaths. The case-fatality rate in Mali was 4.08% and the attack rate in Cape Verde was 2587 cases per 100,000 inhabitants. In Nigeria, Senegal, Mauritania and Mali, the epidemic curves supported by the evolution rates showed an increase in confirmed cases and deaths of COVID-19 during December 2020 and January 2021 compared to the last two months. The effective reproduction rates (R_e_) inferred a slowdown in virus transmission (R_e_ < 1) in these countries except for Senegal.

**Conclusion:** The results showed that COVID-19 was still circulating in some West African countries in late 2020 and early 2021. By improving the health system and with context-specific public health interventions and vaccination, these countries should effectively control COVID-19.

## INTRODUCTION

Coronavirus disease 2019 (COVID-19) has spread rapidly across all continents, and the number of confirmed cases and deaths has increased worldwide [1]. It is the most devastating pandemic that humanity has experienced in the last ten decades [2]. In fact, no other pandemic has taken so many lives in the same time period [2].

Africa confirmed its first case of COVID-19 in Egypt on 14 February 2020 [3]. On 27 February 2020, Nigeria reported the first official case of severe acute respiratory syndrome coronavirus 2 (SARS-CoV-2) in the West African region [4]. On 18 March 2020, Burkina Faso reported the first death [5]. Most West African countries were quick to take measures to limit gatherings and build public health capacity. As a result, the number of cases and deaths has remained lower than in other parts of the world [6-8] despite dire predictions [9]. As stated in the World Health Organization’s draft guidance document on infection prevention and control, it was imperative that member countries strictly adhere to standard precautions for all infected persons, and it was also recommended that a coordinating center for infection prevention and control be established, with the support of senior country and institutional leadership [10,11].

Despite the relatively low spread of COVID-19 in African countries [2], there was a marked increase in cases between December 2020 and January 2021 [12]. On average, 25,223 cases were reported daily between 28 December 2020 and 10 January 2021 in Africa, almost 39% higher than the peak of 18,104 daily cases averaged over two weeks in July 2020 [13]. In West Africa, the first wave had challenged the health system, leading to its saturation. This led Senegal, for example, to opt for home-based management of simple cases [14], which entailed a number of risks, notably the spread of the virus within households and in the community [15]. The most severe ’second wave’ coincided with the emergence of new British and South African variants [16].

Therefore, the estimation of epidemiological measures of coronavirus disease 2019 (COVID-19) is important to assess the extent of epidemic transmission. These results will be useful for policy makers, both at the national and supranational level, to further plan and evaluate measures to contain the spread of the ongoing epidemic. Furthermore, this type of study on the dynamics of the COVID-19 pandemic in West Africa is very limited. The objective is to assess the epidemiological profile of COVID-19 in West African countries from 27 February 2020 (first reported case) to 31 January 2021.

## METHOD

The United Nations defines West Africa as the following 16 countries: Benin, Burkina Faso, Cape Verde, Gambia, Ghana, Guinea, Guinea-Bissau, Ivory Coast, Liberia, Mali, Mauritania, Niger, Nigeria, Senegal, Sierra Leone and Togo [17]. The total population of this region is estimated to be around 381 million people in 2018 [18]. Health is a fundamental issue for West African countries. However, due to their level of development, they do not have health systems adapted to the needs of their populations [19]. This structural deficiency is generally perceptible in the difficulties encountered by states in containing endogenous health threats and combating the major infectious diseases such as HIV and tuberculosis that are rampant in the region [19].

We used the “COVID19R” package, available on the Comprehensive R Archive Network (CRAN), to obtain the data [20]. This package unifies COVID-19 datasets from different sources to simplify the data acquisition process and subsequent analysis [21]. The COVID-19 data are extracted in real time from several reliable sources, including: Johns Hopkins University Center for Systems Science and Engineering (JHU CSSE); World Bank Open Data; World Factbook by CIA; Ministero della Salute, Dipartimento della Protezione Civile; ISTAT - Istituto Nazionale di Statistica; Swiss Federal Statistical Office; Open Government Data Zurich [21].

The epidemiological data for this study included the cumulative number of daily cases of COVID-19 and the daily number of deaths. The study period was from 28 February 2020, the date of the first case detected in the study area (Nigeria) to 31 January 2021. The data covered West Africa as a whole and five countries within it: Nigeria, Senegal, Mauritania, Mali and Cape Verde. These countries were selected on the basis of population (Nigeria), area (Mali), density (inhabitants/km^2^) (Nigeria, Cape Verde and Mauritania), languages spoken (Portuguese, French and English speaking countries) and geographical location (Cape Verde and Senegal) [22].

The analysis was carried out with the R software version 4.0.3 and focused on the following elements:

- The number of confirmed cases of COVID-19
- Cumulative number of COVID-19 deaths
- Case fatality rate by country and globally: proportion of COVID-19-related deaths to the total number of confirmed cases of COVID-19
- Attack rate per 100,000 inhabitants: This is determined by the ratio of the number of confirmed cases of COVID-19 to the total population, multiplied by 100,000
- Epidemic curves by epidemiological week for confirmed cases and deaths of COVID-19: These will allow the trend to be seen globally.
- Effective reproduction rate R_e_: The reproduction rate is defined as the average number of new illnesses caused by an infected person in a population [1]. The effective R is an indicator of the virus transmission dynamics about 1 week before integrating the serial interval. This parameter was estimated using the “EpiEstim” package [23]. We assume that the serial interval distribution follows the gamma distribution with a means of 4.8 and a standard deviation of 2.3 given by Nishiura H et al [24]. For R_e_ > 1, the number of infected patients is likely to increase and for R_e_ < 1, transmission is likely to slow down.
- Rates of change in confirmed cases and deaths: This consists of estimating the change in the epidemic in December 2020 and January 2021 compared to October and November 2020.

## RESULTS

As of 31 January 2021, West Africa had 342,938 confirmed cases of COVID-19 with 4,496 deaths. It had a case-fatality of 1.31% during this period with an attack rate of 89.96 cases per 100,000 inhabitants. Nigeria and Senegal had 131,242 and 26,523 cases respectively (Figure 1).

**Figure 1:**
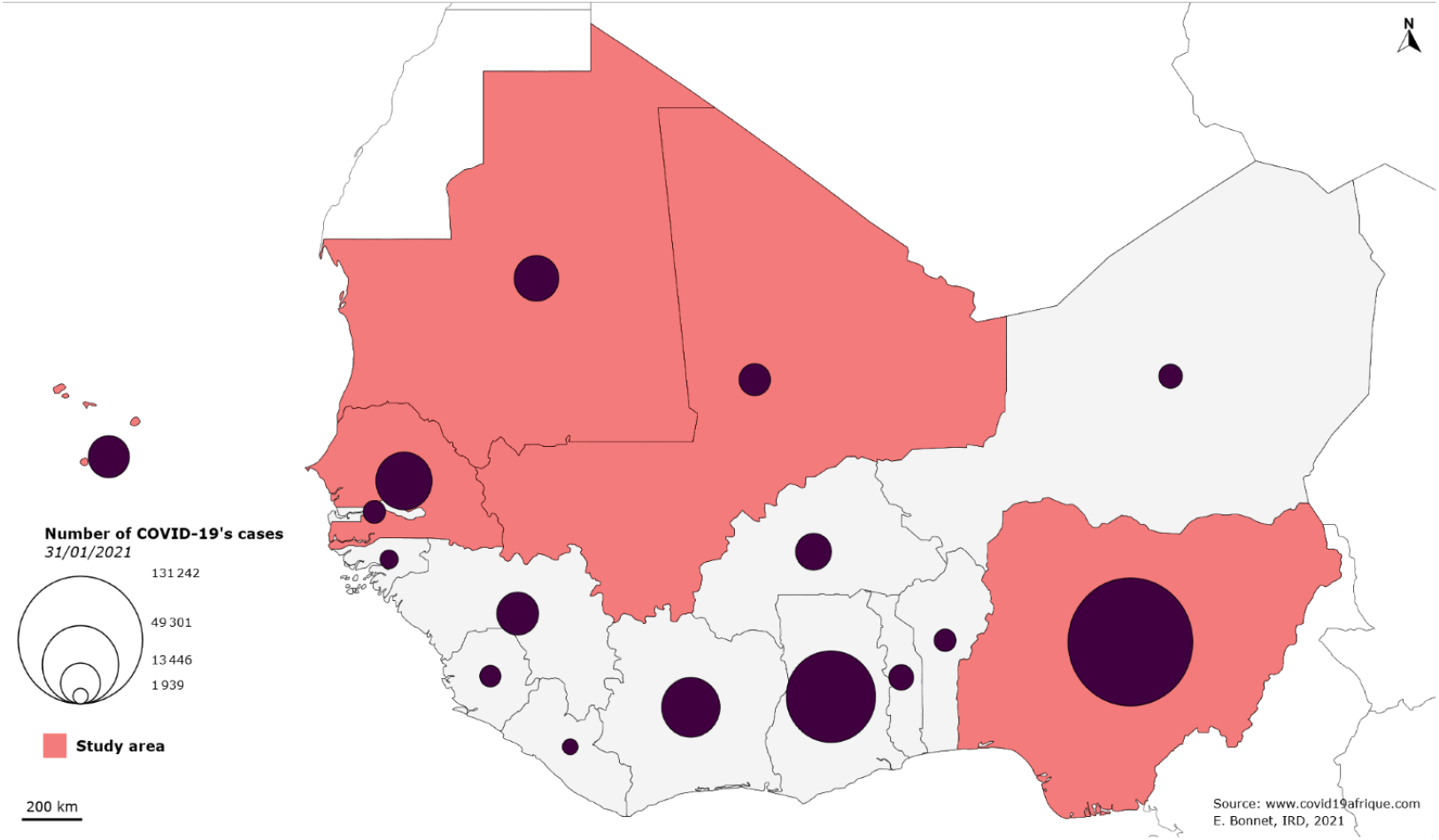
Distribution of the number of cases by country in West Africa to 31 January 2021

Nigeria had the highest number of deaths from COVID-19 with 1,586 deaths followed by Senegal with 628 deaths. Mali had a case fatality rate of 4.08%. The attack rate in Cape Verde was far ahead of all other countries and corresponded to 2,587.51 cases per 100,000 inhabitants (Table 1).

**Table 1:** Status of COVID-19 from 28 February 2020 to 31 January 2021

| Country | Deaths | Population | Case fatality rate (%) | Attack rate per 100,000 inhabitants |
| --- | --- | --- | --- | --- |
| Nigeria | 1586 | 195874740 | 1,21 | 67,00 |
| Senegal | 628 | 15854360 | 2,37 | 167,29 |
| Mauritania | 422 | 4403319 | 2,54 | 377,78 |
| Cabo Verde | 134 | 543767 | 0,95 | 2587,51 |
| Mali | 330 | 19077690 | 4,08 | 42,41 |
| **West Africa** | **4496** | **381196869** | **1,31** | **89,96** |

The epidemic curves of confirmed cases and deaths of COVID-19 showed the same trends (Figure 2).

**Figure 2:**
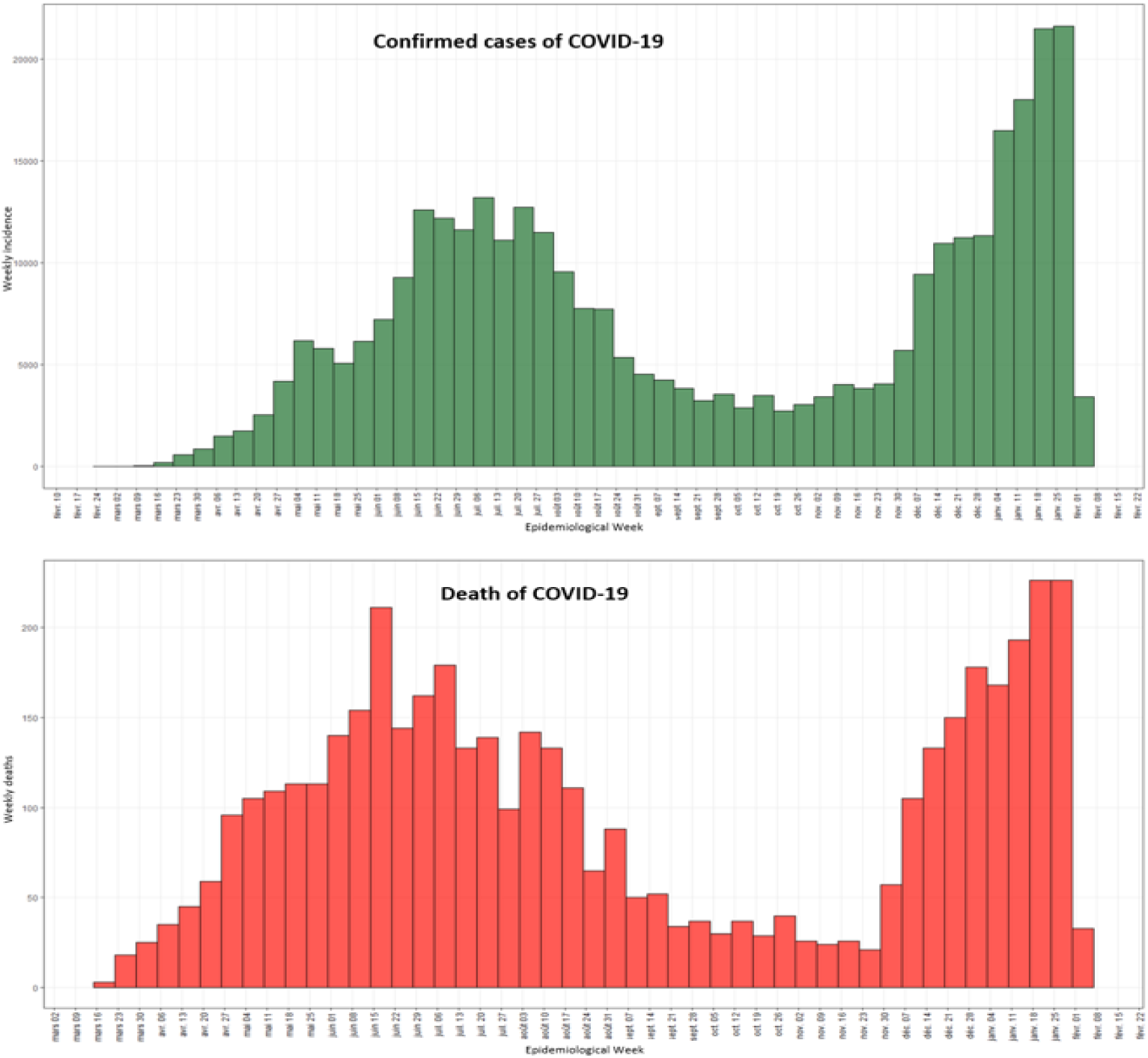
Epidemic curves by epidemiological week for confirmed cases and deaths of COVID-19 in West Africa, 28 February 2020 to 31 January 2021

In Figures 2 and 3, this trend was the same in Nigeria, Senegal, Mauritania and Mali with two waves observed over two different periods. There was a clear trend in confirmed cases and deaths of COVID-19 in these four countries during the period December 2020 to January 2021. For confirmed COVID-19 cases, the rate of change ranged from 842.55% (Senegal) to 112.37% (Mali). For COVID-19 deaths, they ranged from 1431.25% (Mauritania) to 577.05% (Nigeria) (Appendix 1 and 2). The evolution of deaths during the second wave was less important in Nigeria compared to the first “wave” (Figure 3). In addition, the _Re_ in Nigeria, Mauritania and Mali was 0.88 [0.86 - 0.90], 0.77 [0.67 - 0.87] and 0.78 [0.64 - 0.94] respectively, showing a slowdown in virus transmission in these areas (Appendix 1).

**Figure 3:**
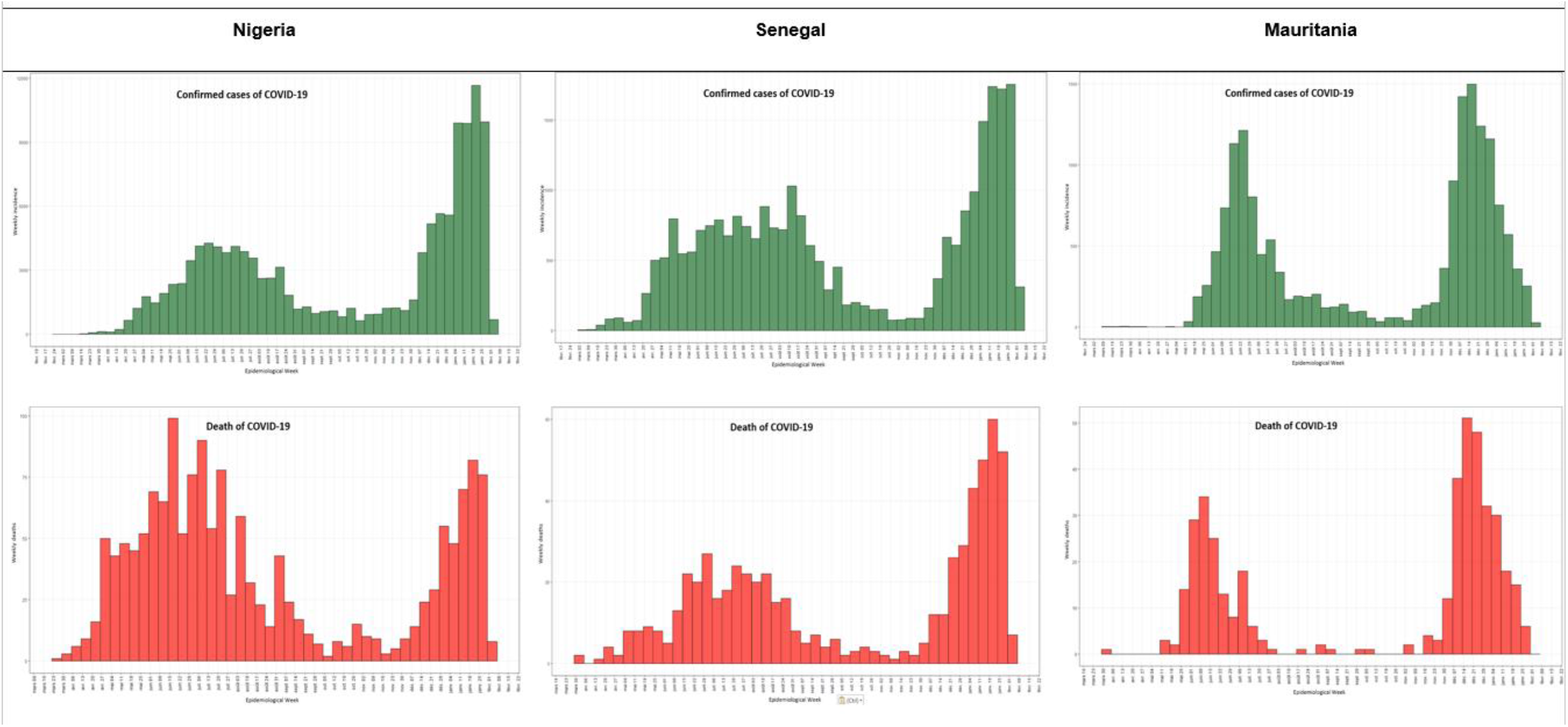
Epidemic curves by week of confirmed cases and deaths of COVID-19 in Nigeria, Senegal and Mauritania, from 28 February 2020 to 31 January 2021

In Cape Verde, this trend was less clear (Figure 4) and there was even a lower evolution of COVID-19 cases and deaths in December 2020 and January 2021 (−30.15% and -35.56% respectively) compared to October and November 2020 (Appendix 1 and 2). However, the _Re_ in Cape Verde was 1.15 [1.05 - 1.25] demonstrating the evolution of the virus transmission in the area during the last week of January 2021 (Appendix 2).

**Figure 4:**
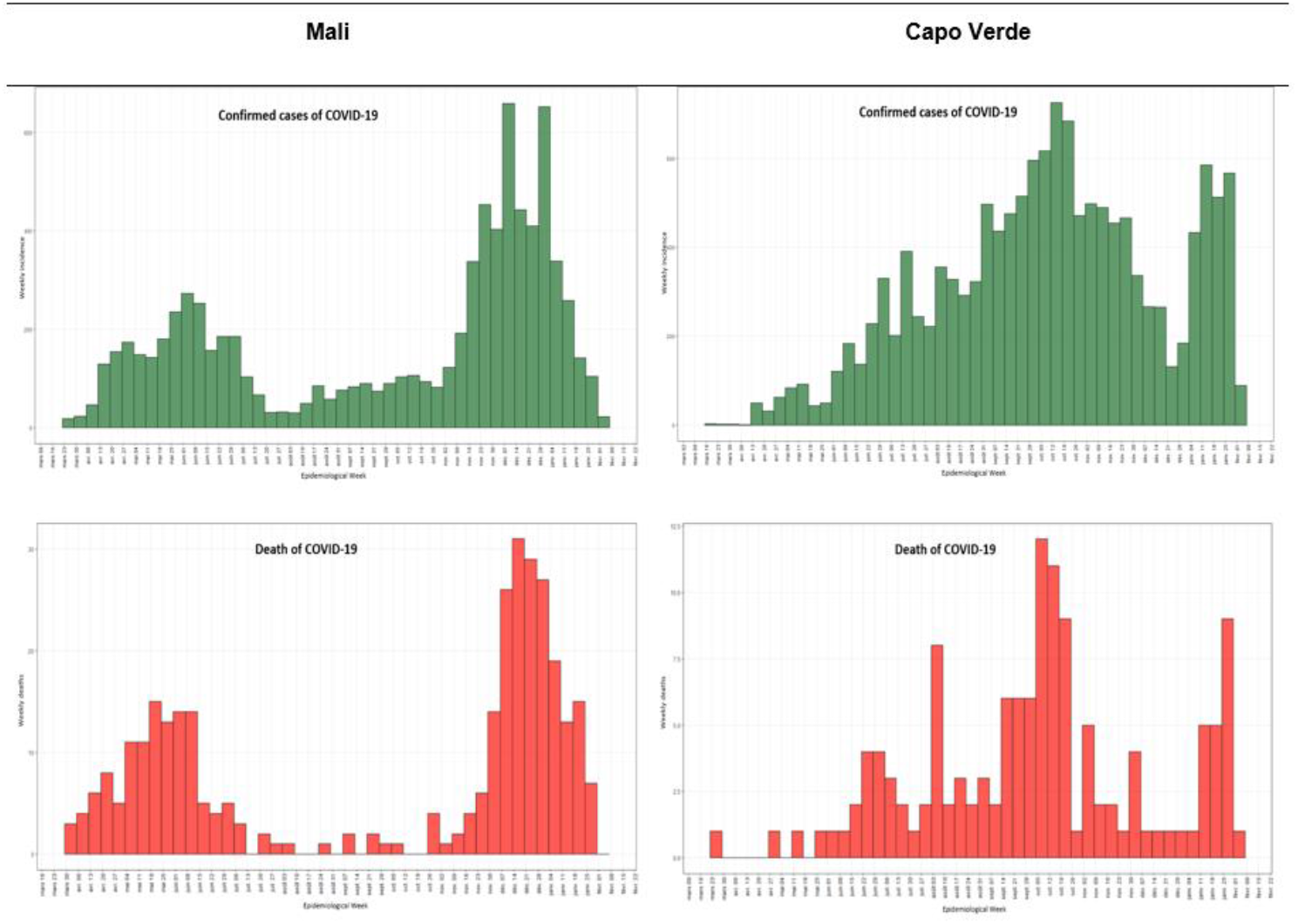
Epidemic curves by week of confirmed cases and deaths of COVID-19 in Mali and Cape Verde, 28 February 2020 to 31 January 2021

## DISCUSSION

This study analyzed the epidemiological profile of COVID-19 in some West African countries. It dealt with the static and dynamic parameters of the epidemic. Globally, transmission of the virus persists, and countries continue to employ various strategies to control the COVID-19 pandemic. In this regard, several countries in West Africa had developed their response plans to elaborate their responses [25]. Thus, they had set up several governmental measures, either before like Nigeria or Burkina Faso [7,26], or coincidentally with the diagnosis of the first national cases. These measures included temperature control in airports, curfews, prohibition of travel between regions, wearing of masks, closure of schools and markets [7,8,26], etc. These different measures did not lead to the development of a national strategy for the prevention of the disease. These various measures did not fully limit the spread of the virus. Although empirical estimates based on the data are not ideal (data irregularities, lack of triangulation with country data, etc.), as of 31 January 2021, West Africa accounted for only 0.3% of confirmed cases and 0.2% of deaths worldwide [27]. Nigeria had the highest number of cases and the highest number of deaths. Nigeria was hit hard by the second wave, which was much larger than the first. It should be noted, however, that Nigeria is the most populous country in West Africa with almost 200 million inhabitants [18]. This may explain why it has the highest number of cases in West Africa.

The case fatality rate of COVID-19 in West Africa was lower than in Africa and the world [27-29]. However, it was highly heterogeneous despite the similarity of health systems, which were characterized by inadequate financing, lack of modern and sophisticated health care facilities, inadequate or non-existent health insurance schemes, shortage of well-trained health personnel, chaotic implementation of health policies and poor access to essential services [2,30]. The causes of this variability presented in this study deserve further investigation.

The number of COVID-19 cases and deaths in West Africa had increased by 326.86% and 451.33% respectively in December 2020 and January 2021 compared to October and November 2020. Nigeria, Senegal, Mauritania and Mali had noted this increase. This could be explained not only by the lifting of restrictive measures that were enacted by government authorities in these countries at the onset of the pandemic [26] but also by the relaxation of preventive hygiene measures [12]. It must be said that West African countries suffered great economic damage during the pandemic [31]. However, trying to revive economies and livelihoods devastated by the pandemic while at the same time aiming to limit the spread of COVID-19 is a difficult balancing act [7,12]. The evolution of COVID-19 cases coincides with the emergence of new variants, although at present this relationship has not been established [16].

Nevertheless, there was a 30.15% and 35.56% decrease in the number of cases and deaths in December 2020 and January 2021 respectively in Cape Verde. The particularity of Cape Verde is that it is an island state. The country had adopted pre-arrival screening in October 2020 after the closure was lifted [32]. This strategy, together with others such as the closure of schools in high transmission localities and the closure of non-essential workplaces [32], could explain the containment of the epidemic in this area. Other hypothetical determinants could explain this decrease during this period, such as the geo-climatic and micro-environmental conditions present on the island [33].

In addition, despite the trend in confirmed cases mentioned above, a decline in transmission was observed in the last week of January in Nigeria, Mauritania and Mali with reproduction rates below 1. This is important and could mean a decline in the transmission trend in the coming weeks. This decline could be reinforced by the vaccination campaign that some have started [34,35]. In particular, it is still important to increase screening, isolate contacts and patients undergoing treatment, and reinforce proven prevention measures.

### Limits

The study does not address the dynamics of the epidemic during the entire 2020 period. It does not statistically compare the magnitude of the evolution of the second wave to the first. Also, it was conducted in only 5 of the 16 countries in West Africa. However, the difference in the context of these countries (geographical location, density, language spoken, etc.) is an important aspect that makes it possible to highlight to some extent the distribution of the health problem in the area. Despite these limitations, the study provides an understanding of the transmission pattern to date and underlines the importance of epidemiological surveillance in countries affected by COVID-19. These data can be used for the planning and implementation of control measures. For this reason, surveillance and simple descriptive analysis should be continued as long as COVID-19 continues to spread.

The data in this article are from 31 January 2021 and in the future the transmission of the disease is supposed to be considered again.

## CONCLUSION

Static and dynamic public health surveillance tools provide a picture of the pandemic’s progress across countries and regions. Although static measures capture data at a specific point in time, such as numbers of cases and deaths, they are less successful in assessing the dynamics of the epidemic. Thus, it was important to incorporate parameters such as the rate of change and the effective reproduction rate. However, the results showed that COVID-19 was still being transmitted in West African countries. By improving the health system and with context-specific public health interventions and vaccination, these countries could effectively control COVID-19.

## Data Availability

Data are available upon request to the authors.

## Data Availability

Data used in the preparation of this article were obtained from the Adolescent Brain Cognitive DevelopmentSM (ABCD) Study (https://abcdstudy.org), held in the NIMH Data Archive (NDA).

## ETHICAL APPROVAL

Ethical approval was not required

## AUTHORS’ CONTRIBUTIONS

MFB and AF: designed the study. MFB: analyzed the data and drafted the manuscript. BD and AF: reviewed the manuscript, and edited the final draft. OB, NMS, EB and VR: provided guidance and comments and reviewed the draft manuscript. All authors have read and agreed to the final version of this manuscript.

---

**Appendix 1:** Dynamics of confirmed COVID-19 cases

| **Country** | **Confirmed cases (October 2020/November 2020)** | **Confirmed cases (December 2020/January 2021)** | **Rate of change (%)** | **R<sub>e</sub> [95% CI]** |
| --- | --- | --- | --- | --- |
| Senegal | 1107 | 10434 | 842,55 | 1,02 [0,97 – 1,07] |
| Nigeria | 8709 | 63685 | 631,26 | 0,88 [0,86 – 0,90] |
| Mauritania | 1099 | 8034 | 631,03 | 0,77 [0,67 – 0,87] |
| Mali | 1592 | 3381 | 112,37 | 0,78 [0,64 – 0,94] |
| Cape Verde | 4737 | 3309 | -30,15 | 1,15 [1,05 – 1,25] |
| **West Africa** | **30101** | **128490** | **326,86** | **1,05 [1,04 – 1,07]** |

**Appendix 2:** COVID-19 death dynamics

| **country** | **Deaths (October 2020/November 2020)** | **Deaths (December 2020/January 2021)** | **Rate of change (%)** |
| --- | --- | --- | --- |
| Senegal | 22 | 295 | 1240,91 |
| Nigeria | 61 | 413 | 577,05 |
| Mauritania | 16 | 245 | 1431,25 |
| Mali | 25 | 174 | 596,00 |
| Cape Verde | 45 | 29 | -35,56 |
| **West Africa** | **263** | **1450** | **451,33** |

